# Navigating the Multiverse: A Hitchhiker’s Guide to Selecting Harmonisation Methods for Multimodal Biomedical Data

**DOI:** 10.1101/2024.03.21.24304655

**Authors:** Murali Aadhitya Magateshvaren Saras, Mithun K. Mitra, Sonika Tyagi

**Affiliations:** IITB-Monash Research Academy, Mumbai, Maharashtra, India, 400076; Indian Institute of Technology Bombay, Mumbai, Maharashtra, India, 400076; School of Translational Medicine, Monash University, Melbourne, Victoria, Australia, 3181; School of Computational Technologies, RMIT University, Melbourne, Victoria, Australia, 3000

**Keywords:** multimodal integration, feature representation, data integration, deep learning, digital health

## Abstract

**Introduction:** The application of machine learning (ML) techniques in classification and prediction tasks has greatly advanced our comprehension of biological systems. There is a notable shift in the trend towards integration methods that specifically target the simultaneous analysis of multiple modes or types of data, showcasing superior results compared to individual analyses. Despite the availability of diverse ML architectures for researchers interested in embracing a multimodal approach, the current literature lacks a comprehensive taxonomy that includes the pros and cons of these methods to guide the entire process. Closing this gap is imperative, necessitating the creation of a robust framework. This framework should not only categorise the diverse ML architectures suitable for multimodal analysis but also offer insights into their respective advantages and limitations. Additionally, such a framework can act as a guide for selecting an appropriate workflow for multimodal analysis. This comprehensive taxonomy would furnish a clear guidance and aid in informed decision-making within the progressively intricate realm of biomedical and clinical data analysis, and is imperative for advancing personalised medicine.

**Objective:** The aims of the work are to comprehensively study and describe the harmonisation processes that are performed and reported in the literature and present a working guide that would enable planning and selecting an appropriate integrative model.

**Results:** We present harmonisation as a dual process of representation and integration, each with multiple methods and categories. The taxonomy of the various representation and integration methods are classified into six broad categories and detailed with the advantages, disadvantages and examples. A guide flowchart that describes the step-by-step processes that are needed to adopt a multimodal approach is also presented along with examples and references.

**Conclusions:** This review provides a thorough taxonomy of methods for harmonising multimodal data and introduces a foundational 10-step guide for newcomers to implement a multimodal workflow.

## Introduction

The growth of biological and healthcare data, in terms of volume, velocity and variety, has been exponential and driven by technological advances in electronics, communication and infrastructure [1, 2]. Concurrently, there has been an increase in data analysis tools to understand and analyse the data. Progress in computational techniques, artificial intelligence (AI) and machine learning (ML) methods have been identified to contribute towards the analysis and interpretation better than traditional analytical methods [3, 4].

Data generated in the context of biological systems can manifest in various forms such as quantitative, qualitative or narrative; each has its subtypes, collectively referred to as a ‘modality’. These diverse modalities can capture several aspects of a biological system, such as nucleic acid and protein sequences [5], gene expression [6], and the biomolecular structure and its activity [7]. Other modalities include the epigenetic state and methylation information [8] of the genome, metabolites, and anatomic and phenotypic data.

Each data type has driven research towards elucidating the corresponding functional aspects to understand the system. Numerous studies using a single data modality have presented valuable additions to the literature in disease mapping, pathway and network elucidation [9, 10, 11]. However, a vast portion of the biological complexity still requires an explanation, an ongoing challenge for the research community.

Different modalities capture different aspects of the system. Thus, integrating them provides a comprehensive multi-view understanding of both biological and clinical conditions [4, 12]. Combining multiple types of omics data or a ‘multiomics’ approach to study biological systems has gained momentum lately due to their demonstrated superiority over single-omics approaches [3, 13, 14, 15, 16]. Furthermore, healthcare data is integrated with omics datasets to reveal their interconnections, providing a comprehensive 360-degree view of an individual’s condition [17, 18]. Such studies have reported significantly validated and reliable results compared to independent analyses. Thus, integrating multiple modalities can reveal synergistic effects, where the combined information enhances the model’s overall performance beyond what individual modalities can achieve.

However, analysing multiple modalities together is challenging, given the heterogeneity of the data. Current research covers the merits of using general ML methods for the analysis of biological and clinical data [19, 17, 20]. It is easier for experienced researchers in the domain to understand and implement hybrid methods and complex ML models. Still, the absence of adequate algorithmic information impedes interested researchers from fully grasping the process and implementing a workflow. Previous review articles on this subject relay information to mitigate data challenges but lack information on multimodal implementation [3, 21, 19, 22]. A definitive explanation of the methods involved in a multimodal approach is missing.

Additionally, A gap persists in delineating between ‘integrated learning’ and ‘co-learning’ methods. While they are used synonymously, they indicate different concepts. Integrated learning refers to a broad, independent analysis of multiple modalities and linking the results to obtain a high-level overview. Co-learning, which we refer to as harmonisation in the current review, allows cross-talk between elements across modalities, enabling to understand the relationships between modalities. Harmonisation aims to elucidate the low-level relationship between features of different modalities [15]. Articles incorporating ‘integration’ as part of their pipeline do not necessarily perform a harmonisation process. Instead, they focus on the correlation between individual data type analysis [13]. The effect of an analysis using multiple modalities is not adequately captured by methods that do not harmonise the features. A co-learning set-up is distinct from an individual analysis since it necessitates a fusion of features.

In summary, this literature review addresses the aforementioned gaps by offering insights into data modalities, challenges encountered in data, the components involved in a multimodal workflow, and a beginner’s guide to multimodal analysis.

## Methods

This systematic review was performed based on the standards of the Preferred Reporting Items for Systematic Reviews and Meta-analyses (PRISMA 2020) statement [23].

Based on existing literature reviews published over the past decade, a general outline was followed to select articles that mentioned multimodal learning techniques. An extensive search of various ML methods focused on biomedical data was initially gathered using the metapub (https://github.com/metapub/metapub) python module based on the following keywords:

*‘Multimodal’, ‘Machine learning’, ‘Integrated learning’, ‘Multiomics’, ‘Genomics’, ‘Proteomics’, ‘Biomedical’, ‘Healthcare data’, ‘Biological network’, ‘Deep learning’, ‘Multitask learning’, ‘Data fusion’, ‘Representation’, ‘Interpretable model’, ‘Neural network’*.

The title and the abstract of the search results reported by the keywords using the python script was used to select papers for complete reading. A few suggested articles from co-authors on were used to initiate the search for reviews on biomedical multimodal integration. Citations from suggested papers were manual searched using Google Scholar and PubMed. Connected Papers (https://www.connectedpapers.com) was used to identify related articles. We identified less than 10 articles that described on the topic. Few papers were selected for reading on general multimodal integration methods. Figure 2 describes the count of articles.

**Fig. 1.**
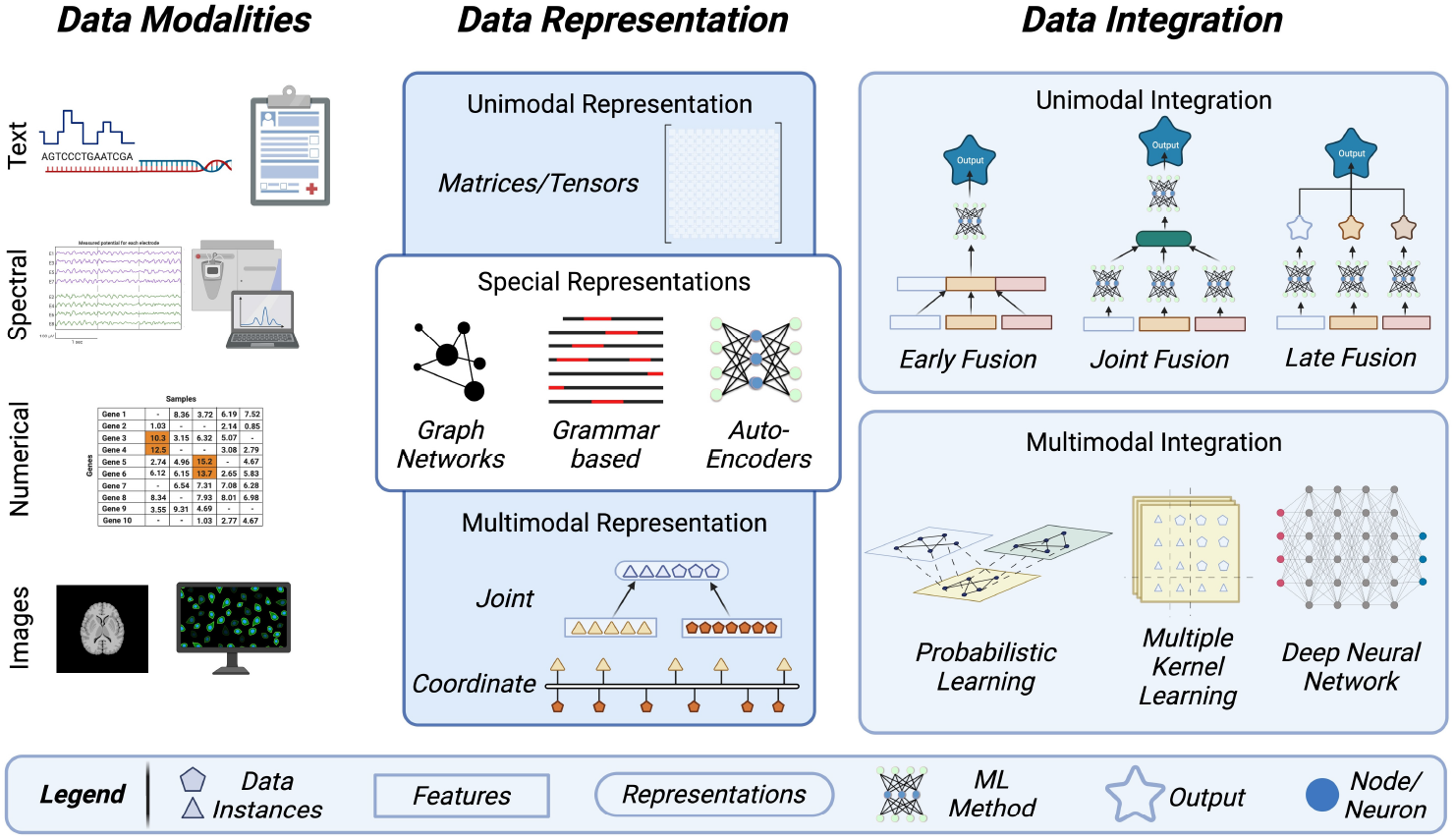
An illustration briefly depicting the broad categories of data modalities and the representation and integration methods used in a multimodal harmonisation analysis. The representation methods are split into three groups based on the number and type of datasets. The integration methods are split based on the type of fusion performed. Made with BioRender.

**Fig. 2.**
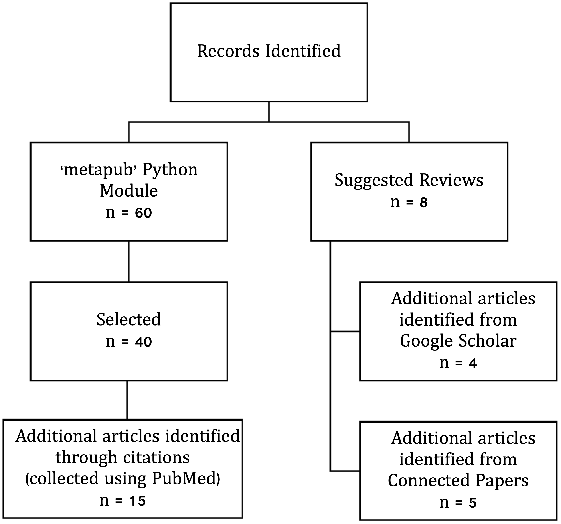
Flowchart of literature screening. The blocks on the left indicate articles searched for representation and integration methods. The blocks on the right describe the targeted search for review articles on biomedical multimodal harmonisation.

The inclusion criteria for this review primarily focused on studies that incorporated multiple different types of biological and medical data towards a singular analysis using machine learning algorithms. Reports that did not use biomedical datasets but employed a multimodal approach for data analysis were also included for review. The exclusion criteria was marked by the absence of a multimodal approach only. However, studies that focused on representation methods of different data types were included for full-text reading. The last date of article search and selection was 15 December 2024.

The methods for selected articles were reviewed in detail and information on the data type, machine learning framework, model advantages, research gaps were collected. They have been classified into groups and presented in tabular format (Tables 1, 4) and the results are discussed in following sections.

**Table 1.**
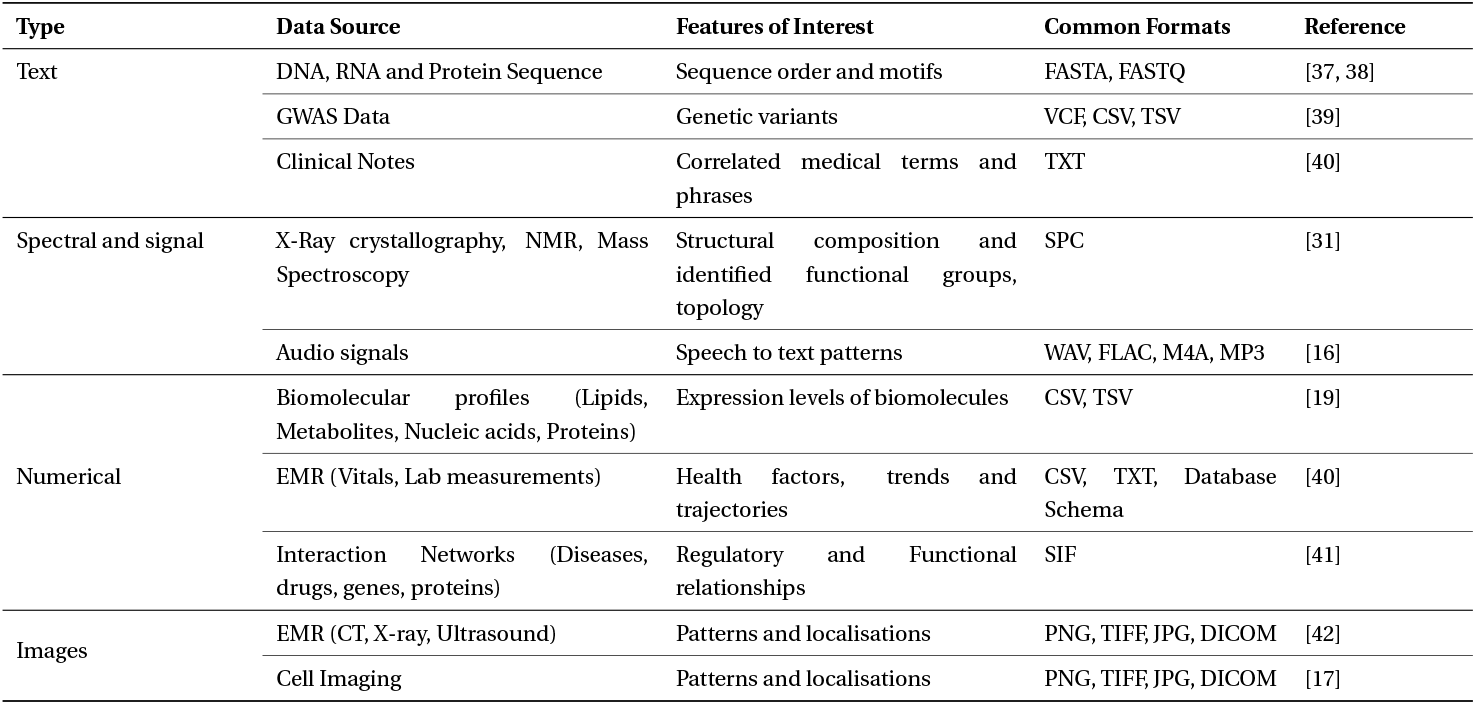
The four distinct modalities biological and clinical data investigated in this study are listed and categorised based on their sources and the features targeted for modelling.

## Data Modalities

In the following sections, we describe the various data types, technical hurdles, and harmonisable methodologies gleaned from the literature we have examined.

### Typical Study Designs in biological and clinical studies

All data generated are usually based on a study or an objective with a focused rationale. The richness of information in collected or generated data depends on its type, determining the range of possible analyses [24].

A static study design acquires data as a ‘snapshot’, that is, collected at a point in time. Case series refer to static data collected from positive-group criteria within a population subgroup, while case-control studies include a negative dataset (controls) for comparison. On the other hand, cohort studies and randomised controlled trials sample data over a period of time, capturing the dynamic nature of a biological system, which allows for a realistic investigation. However, they are resource-intensive methods that must be maintained regularly, and constant follow-up with the subjects considered in the study is crucial. A single type of data is commonly collected from a sample population. Some datasets containing different types of data are generated from the same set of samples, and this is referred to as ‘multi-view data’. Multi-view data offers higher association in inferring correlations between data types because the same samples are used [25].

Many efforts are being taken to enhance the data collection methods and accessibility across various domains, such as in cancer (The Cancer Genome Atlas, https://www.cancer.gov/ccg/) and preterm birth [26]. Current literature predominantly reports on results based on single, static datasets. Correlation studies use multiple datasets to support conclusions through overlapping results [27]. Only a few methods take a complementary approach between modalities [13, 24, 28].

### Common data modalities studied in the literature

Data can be collected in forms such as text, numbers, and multimedia. Based on the sources, they can be classified as ‘biological’ data and ‘health’ data [2]. We refer to ‘biological data’ as information from high-throughput experiments such as sequencing, expression profiling, microscopic imaging and the vast literature corpus for functional annotation. It also includes metadata related to samples, experimental design, assay protocols and technologies. ‘Health’ sources refer to data primarily collected through healthcare providers in digital forms. This data contains an individual’s valuable health and medical history and is stored as time-stamped electronic medical records (EMR). EMR data displays significant structural diversity since it can be structured data, including vital signs and pathology measurements organised in tabular formats, or presented as unstructured data, consisting of clinical notes, images, and documents. Table 1 describes the different modalities that stem from clinical and biomedical sources.

#### Text Modality

Text as a modality comprises various types, encompassing narrative and sequence forms of data. Sequence data stemming from biological macromolecules such as DNA, RNA, and proteins describe and define the relationship between the genotype and the phenotype of an organism. Differences in sequences among groups differing in demography or phenotype are represented as single nucleotide polymorphism (SNP) or small insertions and deletions of DNA bases and are generated by genome-wide association studies (GWAS) [29]. Information on motifs, interaction networks, and annotations about biomolecules, drugs, and diseases belong to this class. Healthcare data, such as EMR, contain unstructured clinical notes and prescriptions manually entered by medical practitioners, which are included in the text category of datasets [30].

#### Spectral and Signal Modality

Spectral data, typically acquired through mass spectrometry experiments to study protein molecules and metabolites, provides detailed insights into the structural composition, constitution and organisation of the molecules under investigation [10, 31]. ML analysis of spectral data involves features representing three-dimensional conformations and spatial relationships of molecules, enabling classification based on functional groups and elements [31, 32]. Proteomics, examining proteins through expression, functional relationships, and structural information, includes investigations into protein folding and structural orientations using methods such as NMR and X-ray crystallography [33]. Structural metabolomics stores the structural data collected from metabolites. Healthcare data in spectral form includes time-dependent electroencephalogram (EEG) and electrocardiogram (ECG) analysed with signal processing methods [34, 35]. Audio data, such as voice notes, undergoes analysis using appropriate methods after feature extraction [36].

#### Numerical Modality

A numerical form of biological data can be from any quantifiable assay, broadly called ‘omics’ data. Transcriptomics represents digital counts of identified expressed transcript molecules, available as a two-dimensional (2D) matrix of genes/transcripts and samples. Similarly, proteomic, lipidomic and metabolomic counts data portray the expression levels of proteins, lipids or metabolite molecules as a 2D data matrix. This modal information allows understanding individual differences regarding genetic expression and linking related biological pathways. EMR readings document vital and pathological parameters such as heart rate, weight, height, age, and blood pressure as numerical time-series data. Frequent time-stamped EMRs enable longitudinal analysis, capturing changes in the recorded values over time [43].

#### Image Modality

This modality encompasses visual information, including images and videos. Videos are also considered under this modality because each frame can be considered an image for processing but in a time-dependent manner [36]. Microscopy and cell imaging data are often analysed for morphology studies, protein localisation and DNA tagging [44]. Manual image analysis methods include segmentation tasks to identify regions of interest and cell morphology assessment [45]. Cell movement and tracking studies creating animated clips from multiple fluorescence-tagged cell images are a modality of this category. Additionally, X-rays, CT and MRI images from EMRs supporting non-invasive diagnosis fall in this category.

### Common challenges associated with biomedical data

Datasets require extensive preprocessing due to incompleteness and imperfections before analysis [46]. Key challenges include high dimensionality, heterogeneity, missing data, class imbalances, bias and accessibility.

Complex high-dimensional data, characterised by large features and file sizes, require extensive computational resources for understanding the variables [47]. Addressing the ‘p*>>*n’ problem, considering the ratio of available samples (n) to features (p), is crucial to prevent specific features from being overlooked in small sample groups [15, 29, 47].

Bias refers to a variety of imbalances found within a dataset and can lead to an unfair interpretation of results. Bias can manifest in various forms, such as representation bias or class imbalance, measurement bias (due to incorrect or unrelated values), aggregation bias (when models are applied to new datasets with a mutually exclusive relationship to training samples) and evaluation bias (when generic models are used as benchmarks for targeted datasets) [48]. Comparative analytical methods utilise representative datasets, subsets of the population with samples from distinct groups like case and control. It is crucial for groups to have samples in a comparable and an equivalent number to understand the true differences.

Irregular clinical data collection processes lead to inconsistent data entries and contribute to missing data. In datasets, all samples may not provide data for all possible features, and the resulting matrix could be sparse in a few cases. Notably, a missing measurement may carry meaning and should be considered subjectively.

Heterogeneity refers to the variety that exists within and across modalities. Within a modality, the data collected across variables can vary in terms of scale, distribution and recorded value, such as discrete, continuous, categories and intervals, due to non-standardised procedures. For example, clinical and genomic data can not be directly compared and analysed, requiring methods to address heterogeneity.

These challenges obstruct the potential in any analysis, but the problem is exacerbated when multiple datasets are involved in a multimodal set-up [49]. Multimodal methods are affected by coherence between dataset sets (due to heterogeneity and missing data), accessibility and computational resources (due to high-dimensional datasets and bias).

Data preprocessing allows one to check, sort, and select data points so that informed decisions can be taken to handle samples with anomalies and poor quality. The lack of data standardisation between multiple collection sites poses a challenge for seamless data harmonisation [50] and requires specific preprocessing for different sources. While imputation methods partially handle missing data, they are not universal solutions, as approximations may not accurately reflect the system [51]. Hence, more data-driven approaches may be adopted in different scenarios [50]. Large datasets can be converted to latent values to reduce computational load. Representation methods effectively resolve these issues.

Biomedical and health data containing personal and sensitive information are restricted for global access, which limits the extent and scope of analysis. Implementing ethical and legal data practices, both nationally and internationally, is crucial for easing the data sharing process [52]. These practices establish a structured and transparent approach to handling data, creating an environment conducive to sharing valuable information.

### Multimodal Data Harmonisation: an investigation of the taxonomy

Multimodal analytical methods aim to combine information from multiple modalities towards one or many of the following goals: 1) Explain a biological phenomenon or phenotype through overlapping results. 2) Account for and impute missing data in one modality through another linked dataset/modality. 3) Condense the high-dimensional, sparse and noisy data to a low-dimensional latent representation.

The fundamental difference between an unimodal and a multimodal analysis is the number of different modalities used. The complete dataset can be directly fed into an ML architecture for an unimodal analysis, but a multimodal approach requires the fusion of features from multiple datasets. The process of merging and modelling features can be classified under ‘representation’ and ‘integration’. The choice of method varies depending on the task to be achieved and the dataset combinations.

### Data Representation

As discussed, biomedical and health data is generated in many forms (Table 1). Data representation methods are crucial as they transform diverse data types into machine-processable formats such as vectors, matrices, or tensors. Vectors are one-dimensional representations of numerical values, while matrices and tensors hold data in 2-dimensional and multidimensional scales. These methods keep track of relationships between elements of each modality via predefined rules, facilitate feature extraction by using relevant information, and help map data from one modality to another. Importantly, the representation methods can be modified to suit the study conducted [20]. In this context, we have classified three groups of data representation approaches.

#### Unimodal Data Representation

Unimodal data representation involves using a single mode or source of information to represent features. Each modality qualifies as an unimodal representation when independently transformed into a numerical format through an encoding or embedding approach.

Encoding involves converting original data into a numerical format, whereas embedding refers to portraying the original data in a vector space that incorporates semantic information. The information from biological sequence and text can be encoded by converting them to a numerical representation based on composition (tallying frequency of words/monomers), K-mers (segmenting a biological sequence as a window of ‘k’ letters) and distribution (percentage of occurrence of each monomer within user-defined ranges) of the sequence [53]. K-mers serve as the counterparts to n-grams or tokens in NLP methods, and ongoing efforts are focused on developing more advanced, data-driven approaches to derive them from sequential data [54]. Many tools have been created to embed text data into a numerical representation, such as word2vec [55] or doc2vec [56], which preserves the order of information and local neighbouring relationships.

Numerical data obtained as-is or in other formats is commonly represented as matrices for analysis purposes. Time-series information is represented as tensors, where the data is nested within matrices, extending in dimensions to include the temporal relationship [57]. Similarly, image data is converted to a matrix representation by splitting a digital pixel into a numerical value between 0-255 for constituent colours. Further, spectral information from biological mass spectrometry studies generates coordinate data and is represented as a matrix.

Unimodal representation is the fundamental way to proceed with any ML analysis. The complete set of features obtained through representation methods can vary in size and dimension depending on the dataset. To alleviate the computational load and resources during modelling, feature selection and feature reduction methods reduce the representation into a smaller latent space portraying the complete dataset, which is used for analysis. There are multiple feature reduction methods, such as the Principal Component Analysis (PCA), Joint Non-negative Matrix Factorisation (Joint NMF) and Autoencoders. Wrapper methods (forward, backwards, and stepwise selection), Filter methods (ANOVA, Pearson correlation, variance thresholding), and embedded methods (Lasso, Ridge, Decision Tree) are all part of feature selection techniques [22].

#### Multimodal Representation

Multimodal data representation involves using multiple modes or sources of information to represent data. A multimodal representation fuses multiple unimodal representations together onto a shared feature space (joint) or corepresents the features from the different datasets (coordinate).

Each modality is condensed in a joint multimodal representation, and the defining features selected are concatenated to form a single collective representation. The ratio of features from each modality contributing to the concatenated representation is maintained uniformly. This prevents modalities with fewer features from being overwhelmed by modalities with large dimensions. Zhao et al. describe the application of joint representation in two publications using image data and clinical information [58, 42]. They merge representations of image data (CT scans) and clinical information in different ratios and predict lymph node (LN) metastasis [58]. In a subsequent publication focusing on the same diagnosis, they introduce the 3M-CN architecture that utilises a ‘refine layer’ to predict LN metastasis [42]. The refine layer is a concatenation of key features identified from clinical information and processed 3D images.

Coordinated representations reduce and present the features within each modality individually but link them towards the same meaning over a common coordinate space. Trajectory inference or pseudotemporal ordering is a method to classify the different stages of the same cell type along an axis representing evolution [59]. Pseudotime ordering is an excellent example of coordinate representations, where data from single-cell experiments are projected onto an evolutionary axis [59]. The relationships established with identified patterns and domain knowledge help associate the features. MATCHER is a tool that has depicted imputation and correlation between modalities using a coordinate representation [28]. The manifold alignment method used in this tool achieved this task by representing data in low dimensions called a manifold and aligning them in a common space (alignment) [60].

Multimodal representation methods empower ML architectures to investigate the interplay between features across diverse modalities. The entities in a biological system interact with each other in varied ways. Hence, the ratio of representations in the shared space as a parameter also affects the results of a multimodal analysis [58]. Coordinate representations become more complex than joint representations when there is no common ground to link the features.

#### Special representations

Special approaches represent data non-conventionally through a generative or rule-based approach. These are not mutually exclusive to the previous two categories but process one or more source modalities differently to generate a representation. Generative representations learn the underlying patterns and structure of the data and are capable of generating new instances of data that are similar to the examples they were trained on. On the other hand, rule-based representations leverage formal rules and semantics to describe the features within a dataset.

### Auto-Encoders (AE)

AE methods compress the entire dataset into a compact set of dimensions through an ‘encoding’ process, eliminating any non-representative features. A ‘decoding’ process then reconstructs the original data using the condensed representation, validating the reduced feature space. The decoding layers are generative of the relationships between all the variables within the data, and hence, this method is classified under a generative representation approach.

Detlefsen et al. extensively explores AE-based representations, emphasising the superior results achieved through the non-linear representation method in various tasks [61]. Zhang et al. introduce OmiEmbed as a multitasking framework, utilising the low-dimensional latent space generated by AEs for downstream tasks like cancer classification and survival prediction [62]. AE representations also find applications in gene identification and cancer detection using expression data [63] and predicting carcinoma primary sites through DNA methylation data [64].

The encoding process in AEs can incorporate any type of model, such as a fully connected neural network (FCNN) or convolutional neural network (CNN), to generate the latent space [62]. Multiple modalities can also be combined at the input to generate a joint latent space [65]. This allows to generate different variants of the latent space and increases the choices available to work with. The validation by the decoding process makes the latent space devoid of errors or data misrepresentation. However, the interpretability of AEs is generally low, and reducing the dimensions of the latent layer further diminishes the model understanding.

### Graph-based

Graph representations portray relationships between different biological entities as a network by considering all features as ‘nodes’, and the relationship is depicted using ‘edges’. The edge values denote characteristics like similarity, interaction, and affinity between nodes based on the data. Graphs can be generated from experimental data (e.g. omics), theory and literature (e.g. disease networks) to represent qualitative and quantitative information. Clinical information about diseases and drugs can also be represented as graphs with links depicting common pathologies and targets. Intramodal networks describe relationships between identical node types (for example, protein-protein), while intermodal networks depict links between distinct node types [41]. Omics modalities such as genome, lipidome, metabolome, proteome, and transcriptome can be fused with environment and EMR modalities and represented as a Heterogeneous Multi-layer Network (HMLN) [66]. Sparse data matrices are easily translatable into graphs, as they efficiently condense large-dimensional data to relevant nodes.

Specific ML architectures are devised to best use a graph network representation to map and predict links between nodes. Graph Convolutional Networks (GCN) create features from local graph structures to learn, and they can be scaled based on the number of interactions to represent complex relationships [67]. Graph Attention Networks (GAT) incorporate attention mechanisms to overcome structural overfitting for higher-order GCNs [68]. Ghorbani et al. presents the MGCN architecture, which implements graph representations to consolidate multilevel data [69]. The methods are highly sensitive to missing and unseen information but excel at discovering links within datasets.

### Grammar-based

Grammar-based or semantic methods rely on a predetermined, ordered set of ‘vocabulary’ to generate a representation, usually for a text-based modality. High-level patterns observed in the modality are identified, and the complete dataset is represented based on the patterns discovered through a feature generation procedure. Additionally, data can either be embedded based on the provided dataset or referenced from the complete knowledge bank. Dictionary-based embedding methods create embeddings for the complete corpus, and the available data is represented based on the closest relationship from the complete dictionary [15].

Tyagi et al. used grammar-based representations to model the syntactic and semantic rule of RNA folding and used context-free grammars (CFG) to generate sequences and parse their structures [37]. Andikos et al. created Knotify [38], a tool to predict RNA pseudoknots using CFG. Onokpasa et al. assert that CFG representations improve compression ratios of RNA sequences and structures [70]. Grammar-based representations have been used to embed the motif information from sequences with the domain knowledge to depict the functionally connected regulatory regions [71].

Although grammar-based representations are powerful in capturing structured information, they may face challenges in handling the inherent ambiguity and variability in real-world data. They require large amounts of data and computing power to process and generate the rule-based representation [15].

In a few cases, ML methods do not differentiate between the representation steps and model training. For instance, dense neural networks and deep-learning architectures do not explicitly have a joint representation stage. They are directly processed to learn the features of the data and train the model (Section DNN).

### Fusion of Data Modalities: Integrating Multifaceted Information

Data fusion methods harmonise the different data modalities available for tasks such as clustering, regression, or classification, utilising the representations from the methods discussed above. Different modalities can be harmonised using two broad ML approaches: Unimodal learning and Multimodal learning.

#### Unimodal Learning

In a multimodal intergration process, this approach treats each modality individually, and each modality requires dedicated processing before fusion occurs. Here, three categories of unimodal learning-based integration exist based on how the features from different modalities are fused: (i) Early, (ii) Late, and (iii) Joint integration.

##### Early Fusion

Early fusion methods describe ML architectures that concatenate feature representations from multiple modalities at the input stage for modelling. The data is minimally processed, primarily to resolve heterogeneity, and samples are removed if imputation is impossible for missing data. Since the data is fused at the input stage, only one unified model is needed for training and inference. This reduces the complexity of managing multiple models or sub-networks for each modality. This method disregards prior selection bias and allows us to investigate all features across modalities. It is time-consuming and computationally expensive to process the complex combinations of all features from modalities [2].

The benefits of using early integration methods are discussed and reported by Barnum et al. [72]. They assert that using immediate fusion techniques to merge modalities before feeding them into a model works better by integrating the lowest statistical correlations between input features. Banerjee et al. describe the PERFORM algorithm, utilising EMR data represented as temporal vectors, to assess its prediction performance in diagnosing acute pulmonary embolism (PE) [40] with ElasticNet architecture [73].

In the case of early fusion, class imbalance and differing sample sizes across modalities can affect the contribution of individual datasets, potentially biasing the analysis. Moreover, as the number of harmonised modalities increases in early fusion methods, the interpretability of the model drastically reduces. The limited coherence among data modalities restricts their combined usage, creating challenges in achieving a unified representation. For example, input data as a combination of metabolomics and chromatin accessibility data may hinder a unified representation. Chen et al. describe data agnostic and data specific methods, with choices of modalities that can be used for coherent analysis [14].

##### Late Fusion

Late fusion methods analyse multiple modalities independently using a model that best fits its representations to a predicted output, and the outputs from each are aggregated towards a singular result or inference. Late integration is generally performed either by taking an aggregated average of the predicted probabilities (outputs) from each modality or passing all the predictions from each modality into an FCNN to process a final output.

Wang et al. presented MOGONET as a tool to classify cancer subtypes using three modalities: mRNA expression, methylation and miRNA expression [74]. A late fusion architecture was established using GCN to predict an initial class label and an FCNN to generate a final class label prediction. Luo et al. presented a modified version of MOGONET, called GRAMINet with GATs instead of GCNs [75]. Both examples pass the same combination of modalities through different architectures to learn a model for biomedical data classification. However, this is not universally applicable since the late integration methods provide flexible model selection for different modalities. Huang et al. investigated a multimodal approach to predict PE and reported the results on seven different architectures, one early, two joint and four late fusion architectures [76]. The early fusion methods had the highest sensitivity, while the late ElasticNet architecture outperformed in all other metrics, such as accuracy, AUROC, specificity and positive predictive value.

Ensemble learning may be considered a variant of the late fusion model, where the outputs of multiple ML models are combined towards a final decision [77]. Late fusion focuses on combining features or representations after individual processing, whereas ensemble learning leverages the diversity of multiple models to improve overall predictive performance.

Late fusion methods do not directly allow for the interaction of features from multiple modalities. It enables training each modality with independent, unique models without interference from other data types. As a result, concerns about different dataset sizes, heterogeneous measurements, and model compatibility vanish. The ability of late fusion methods to capture all information within each modality equally makes it the widely reported harmonisation method in the literature.

##### Joint Fusion

Joint fusion methods endeavour to extract a representation of all initial modalities and model them together to a predicted output. Direct concatenation methods, often used in early fusion, are not possible between modalities that have different quantities and may require a heavy preprocessing step. In late integration, the interdependency between features across modalities is ignored. Joint integration methods provide an advantage through the interaction of features from different modalities in the training phase, irrespective of the observed heterogeneity. The heterogeneity is mitigated since the feature representation and selection procedures reduce and unify the information numerically.

Joint integration methods have been explored using modalities such as CT scans, EMR, methylation, and expression data to achieve biomedical tasks of classification towards prognosis and diagnosis [58, 42, 74, 76]. Zhao et al. investigated the effect of different ratios of EMR features during an integrated analysis of CT images using the DensePriNet architecture [42]. Huang et al. propose a joint representation of CT images and clinical features in a ‘refine layer’ to predict an output that performs better in detecting pulmonary embolism in comparison to other models [76]. MATCHER utilises a joint fusion method to interpolate instances based on the alignment of multiple modalities to the pseudotime scale [28]. Multi-view datasets are analysed with a joint representation and fusion to obtain linked information between modalities. MOMA integrates two modalities from the same sample set towards cancer classification [78], and iDeepViewLearn presents an algorithm for more than two modalities [79]. DeepIDA-GRU jointly integrates both time series and static data in their algorithm [80].

Joint methods are more complex than late fusion but easier to interpret than early fusion methods. Using representations rather than the raw features from each modality greatly dramatically the computational load compared to early fusion. However, joint harmonisation methods are not data agnostic and may require carefully curated model designs.

### Multimodal Learning or Co-learning

Multimodal learning is designed to integrate and model the features from different modalities more comprehensively than unimodal learning. Joint fusions merge representations, but multimodal learning enables co-learning through a direct feed of all the features interacting at the lowest, complex levels. They can be classified into three distinct co-learning methods: probabilistic, multiple kernel learning and deep neural networks.

#### Probabilistic Models

Probabilistic methods use joint or conditional probabilities to build models that capture the relationships and dependencies between different modalities. These models are highly interpretable, allowing them to integrate expert knowledge in the fusion approach and allowing us to interpret the results better than other methods. The random walk method uses probabilistic values to simulate a particle moving between nodes and layers in a network, establishing the relationships and links between the nodes [81].

MultiXRank module, published by Baptista et al., is an example of the probabilistic method of integration using a multiplex network of intramodal and intermodal interactions (protein-protein interactions, gene multiplex and disease monoplex networks) [81]. Pio-Lopez et al. describe a use case of the random walk with restart architecture, wherein the method predicts long-distance gene-disease interactions using gene interaction network and disease similarity network data [82].

Probabilistic methods are applicable to any combination of modalities as long as they form a multiplex network. They rely heavily on theoretical knowledge to bridge relationships between elements of multiple domains and hence can be applied to data from any domain with multiplex and bipartite networks [82].

#### Multiple Kernel Learning (MKL)

Kernels are linear classifiers that divide the data linearly using lenient boundaries, and a combined multitude of them assist in classifying non-linear heterogeneous data [83]. This method is implemented in support vector machines (SVM), a popular method to analyse complex data.

Liu et al. used SVM to model MRI datasets from multiple sources towards Alzheimer’s disease classification [84]. Lancktiet et al. predict the functions of yeast proteins using kernel-based learning [85]. Multiple matrices describing the protein data were used in the algorithm, and results were reported on the different combinations of kernels used to classify the proteins as per their functions [85].

In MKL, different kernels are applied to each modality, and the combination of these kernels is learned to optimise the model’s overall performance. Kernels can identify linear boundaries in datasets, making MKL a highly suitable method for classification tasks [86]. Kernels can be combined in different ways (sum, product) to generate new kernels. A combination of multiple kernels accounts for a better classifier than using a single kernel [87]. This method is resistant to outliers but is susceptible to missing data [86].

#### Deep neural networks (DNNs)

Deep neural integration methods are characterised by a substantial number of neurons and layers constituting neural networks with significant depth and complexity. DNNs utilise representations of different modalities to reduce the features and pass them through high-level, intricate architectures, which enable them to uncover hidden information within the datasets.

DNNs are extensively used to understand data at a microscopic level, especially in the biomedical domain. EMR data can be modelled with omics modalities to shed light on physical and phenotypic changes and their relationships across time. Zhu et al. address an ML model to fuse and learn time-series data, with the use of Stacked Sparse Auto-Encoder (SSAE) and Long Short-Term Memory (LSTM) architecture [57]. Zhang et al. have reported about OmiEmbed, a multitask deep-learning framework based on an autoencoder architecture [62]. AffinityNet, proposed by Ma et al., uses k-nearest neighbours (kNN) attention pooling where the cluster representations of the data are processed as a GAT [88]. The method has asserted good performance for both labelled and unlabelled datasets.

DNNs are computationally expensive to perform due to their dense and complex architecture. They model data at high degrees of non-linearity, but the process becomes hard to decipher and elucidate. The meaning of data is lost when modelling and remains a black box with very low interpretability.

## Guidelines for Model Selection

We propose ten recommendations for initiating a multimodal harmonisation analysis (Figure 3. Articulated objectives and aims are essential before initiating the harmonisation analysis. These objectives will guide subsequent data collection, representation, and model selection steps.

**Fig. 3.**
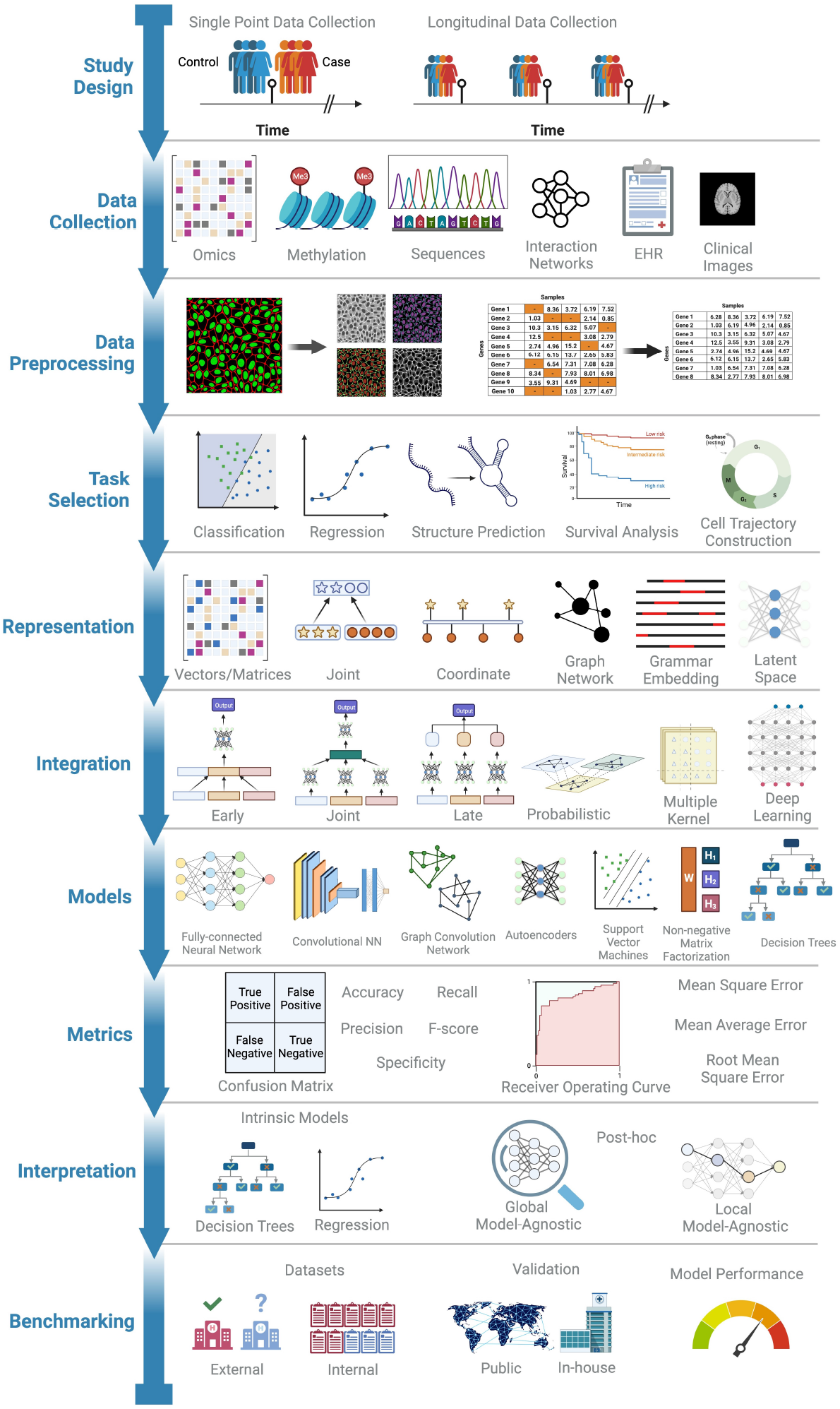
A ten step guide flowchart that describes the process and order of execution to perform a multimodal integration. The titles on the left of the timeline describe the task order. The illustrations on the right are representative examples of different methods under each category. Made with BioRender.

### 1. Tailor Study Design to Objectives

Tailor the study design to the defined objectives, taking into consideration the scale of the study and available resources. Ensure effective study design for sample identification and data collection that aligns with the study’s goals [89, 90].

### 2. Implement Optimised Experimental Protocols

The data is either already available or generated through new experiments. Employ optimised protocols such as the FAIR Principles (Findable, Accessible, Interoperable, Reusable) for data and metadata collection strategies, ensuring consistency and reliability [91]. Here, both the study and software design protocols form the foundation for subsequent analysis steps and contribute to the quality of research outcome [92].

### 3. Interoperable Global Sharing

Digitise all collected data to facilitate analysis and global data sharing through repositories and databanks. Adopting interoperable data and metadata standards enhances data sharing and harmonisation. It is a crucial step for collaborative research efforts and ensures data accessibility for future studies [91, 93].

### 4. Modality Identification and Data Preprocessing

Classify the collected data into the broad modalities discussed here and systematically process the data individually to create a working subset free of artefacts and low-quality elements. Care must be taken during preprocessing to ensure minimum signal loss, and more data-driven approaches as opposed to ad-hoc rules must be used to discard uninformative data points. The analysis and results can vary depending on the level of preprocessing done [94, 50].

### 5. Task Selection

Analysis of the target variable depends on the task selection, and it should be guided by the study’s goals. Choose tasks that reflect the aims and are applicable to the dataset.

### 6. Choose Feature Representation and Integration Methods Wisely

The choice of representation or integration methods influences each other. We recommend selecting them according to the type of data, the number of modalities, the level of harmonisation, and the coherence of the modality. Early and joint learning can be employed for linked modalities and late fusion for dissimilar data types. Recognise that a one-size-fits-all approach is impractical, and tailored methods may be needed for different tasks (Table 2, 3).

**Table 2.** Representation methods detailed with their advantages and disadvantages.

| Representation |  | Advantages | Disadvantages |
| --- | --- | --- | --- |
| Unimodal |  | Simple; Interpretable; Allows interdependencies | Cannot capture contextual information; Susceptible to noise or biases; High feature sizes |
| Multimodal | Joint | Combines features to common space; Controls modality size effects; Allows interdependencies; Interpretable; Reduces dimensions; | Requires tailored architecture; Relies on meaningful cross-modal relationships; |
|  | Coordinate | Aligns features to common space; Controls modality size effects; Allows interdependencies; Interpretable; Reduces dimensions; Imputes information; Captures contextual Information | Requires common axis for representation; representation depends on quality and definition of common space |
| Special | AEs | Creates latent representations; Controls modality size effects; Allows interdependencies; Reduces dimensions; Low susceptibility to noise | Low interpretability; Computationally expensive; |
|  | Graph | Represents qualitative and quantitative; Interpretable; scales with feature size; covers global information | Requires domain-specific knowledge for feature extraction; Complex algorithms needed for irregular structures and dynamic graphs; Susceptible to missing information |
|  | Grammar | Applies for text modality; Captures patterns and semantic information; Reduces dimensions; Interpretable; | Affected by ambiguity and complex language constructs; Computationally expensive; Large datasets needed for processing; Susceptible to missing information |

**Table 3.** A description of the integration methods and their advantages and disadvantages for a multimodal set-up.

| Integration |  | Advantages | Disadvantages |
| --- | --- | --- | --- |
| Unimodal | Early | Requires dedicated processing for each modality separately; Can performs lower-level of statistical correlation between features; Disregards selection bias | Computationally complex and expensive; Prone to data loss as matching features across modalities are required. Requires tailored models for effectively modelling and interpreting features |
|  | Joint | Mitigates heterogeneity during modelling; Reduced feature size due to representations eases computational load | Requires tailored models for modelling joint representations |
|  | Late | Independent models for learning different modalities; Ignores representation bias | Does not allow for feature interaction |
| Multimodal | Probabilistic | Highly interpretable; Establishes links between entities across different modalities | Requires domain-specific multilayer network information; Susceptible to missing data |
|  | Multiple kernel Learning | Models based on overlapping results; Applicable for non-linear relationships; Resistant to outliers; Interpretable | Susceptible to missing data |
|  | DNNs | Uncovers hidden information without explicit rules; Utilises complex architectures to understand the non-linear relationships between features and modalities | Low interpretability; Computationally complex and expensive |

### 7. Navigate Model Selection Complexity

Different ML models can be employed for the same harmonisation set-up. Employ different models of varying complexity to assess the data and evaluate their performance using appropriate metrics (Table 4).

### 8. Model Performance Metrics

Select model performance metrics corresponding to the task to compare and choose the optimal model. Provide an explanation of the metrics used and their relevance to the task [95, 56, 96, 57, 97, 98, 99].

### 9. Prioritise Interpretable Models

Prioritise using interpretable models, either intrinsic or through post-hoc interpretation. Shallow ML methods, such as decision trees and regression fit, explain the relationship between variables, while model-agnostic methods relate the sample to the variables. Especially in clinical settings, understanding how a model arrives at conclusions enhances trust and reliability [100].

### 10. Validate and Benchmark Models

Validate models on different datasets and sources to ensure robustness and generalizability. Benchmark models against state-of-the-art approaches and external datasets to mitigate aggregation and evaluation biases [101, 102].

Overall, these recommendations must be adapted to the specific context and goals of any multimodal harmonisation analysis.

## Discussion

### Lack of Comprehensive Reviews

The article highlights a noticeable gap in the existing literature regarding comprehensive explanations of workflow and procedures for integrating biomedical multimodal data. Multiple reviews for machine learning strategies to process multimodal data are available, but there is a deficit of articles relating them to biological and clinical data. A predominant part of the research literature presents results with information from a single modality. Studies that utilise different biological modalities often interpret the results of independent analyses together. The concept of co-analysis, or more aptly, ‘co-learning’, is missed. There is a lack of clarity on how to effectively integrate data from disparate sources at the lowest item level to extract holistic knowledge.

### Diverse Taxonomies in Multimodal Analysis

Biomedical multimodal data from the same sample set is now routinely available from various research and development activities and healthcare. In the context of multimodal analysis, there is a distinction between the representation and integration steps and unimodal analysis. We highlight the various analysis methods and the data types available under a limited set of taxonomic categorisations.

This classification of data types from biological and clinical sources allows one to identify methods that will suit the analysis of specific combinations and evaluate the advantages of each. We describe data harmonisation as a split of representation and integration methods, each with six distinct categories. Most steps are similar to an unimodal analysis, and the distinction in a multimodal analysis arises in the representation and the integration steps. The representation methods proposed emphasise the features within the data. The various types of representation methods are key to uniformly presenting the multimodal data prior to an analysis. The section on integration focuses on the various methods to feed the data into ML architectures. The article discusses this difference and provides insights into how to handle representation and integration methods for multimodal data effectively.

#### Framework and Model Suggestions for Biomedical Data Combinations

There is a need for a structured framework or guideline to facilitate the harmonisation process for multimodal data. The article addresses this gap by presenting the first guideline framework towards a data harmonisation process and providing a complete workflow. The recommended procedure consists of 10 steps to plan through towards a multimodal analysis.

To assist those undertaking harmonisation for the first time, we present a guide matrix showcasing examples from published literature, illustrating different combinations of data modalities. The combinations between the representation and integration methods are presented as a non-exhaustive list in table 4. Existing studies show that different choices can yield different results when using the same datasets [76]. The diverse taxonomies outlined in this paper can assist in understanding the significance of choosing an appropriate integration model for analysis, considering the concern related to biomedical data and model challenges.

#### Future Focus for Harmonisable Models

The article acknowledges the challenges related to data and model selection in the context of multimodal analysis. It suggests that diverse taxonomies outlined in the paper can assist in understanding the significance of choosing an appropriate integration model for analysis, considering these challenges.

Data-related challenges and model-related challenges both arise when implementing a multimodal analysis. In addition to the challenges related to biomedical data, concerns about data acquisition and maintenance also require attention. The quality of biomedical data collected needs to be maintained, with appropriate measures taken to de-identify the data and global sharing. A vast majority of the published literature on biomedical multimodal analysis focuses on the model metrics and parameter scores. However, due focus should be given to the model interpretability as well. Multimodal analysis with complex architectures may yield high performance scores, but they cannot be used to understand the biological and clinical data if the models are not interpretable. Many of these models offer interpretability-important edges from graph networks, tokens from grammar-based, weights from attention, and ranked kernels are a few examples that can be linked to the data from the model. Interpretable models are needed to understand the process, especially with biomedical data, and to relate to further procedures, such as diagnosis and intervention strategies. Examples provided in Table 4 detail the level of interpretability of different ML models.

**Table 4.**
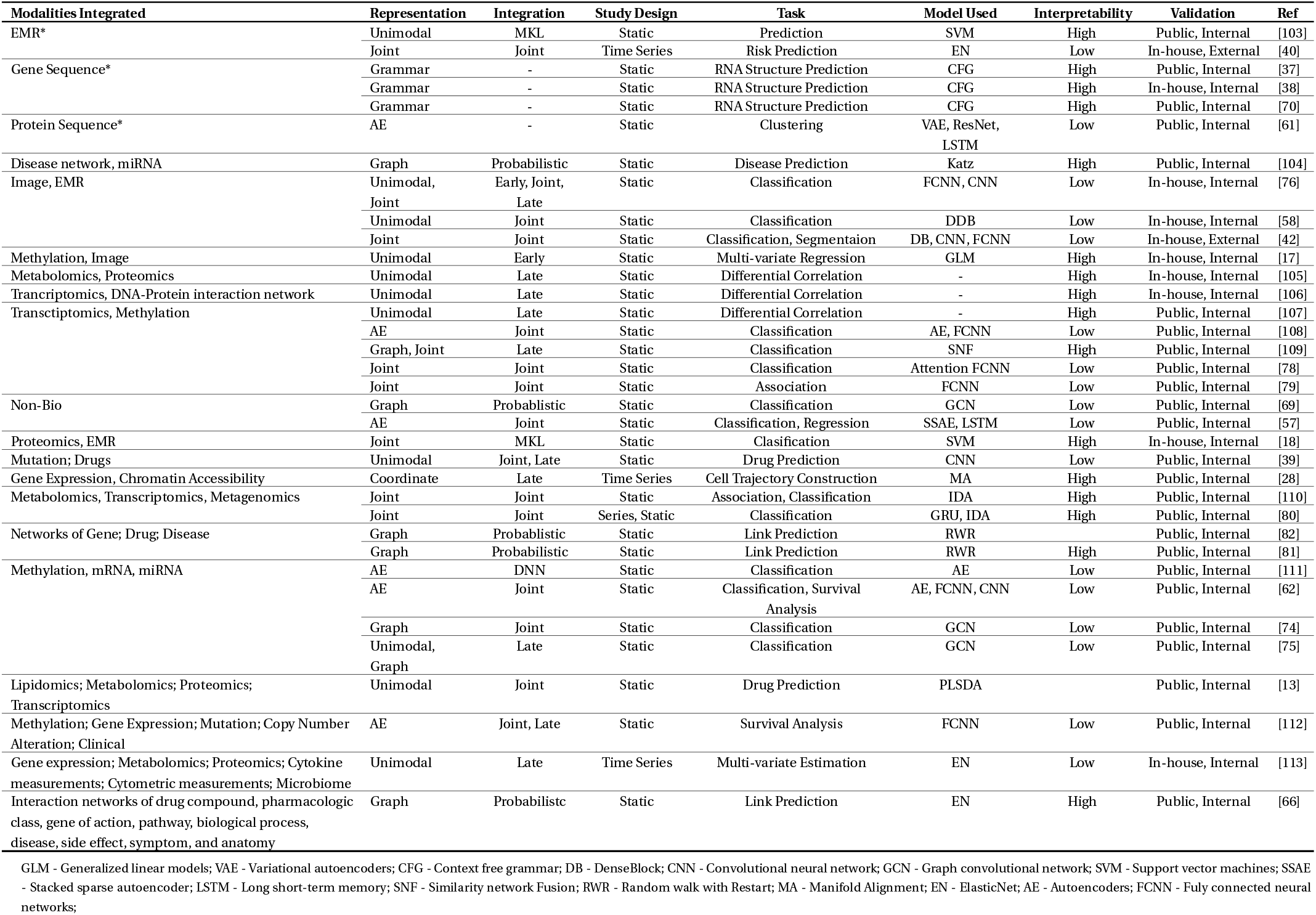
Guide matrix providing examples from literature based on different combinations of modalities and the corresponding tasks, study design, representation, integration, model used, interpretability and reference. * are all representation methods

## Conclusion

The article highlights a significant gap in existing literature regarding the integration of multimodal data, noting a lack of comprehensive explanations and holistic views in current research. While recognising the transformative potential of multimodal integration, it emphasises the need for clarity on effectively integrating disparate data sources to extract comprehensive knowledge. Acknowledging challenges in data and model selection, the article proposes using diverse taxonomies to aid integration model selection. Addressing the distinction between unimodal and multimodal analysis, the article provides insights into representation and integration methods for multimodal data. Furthermore, it underscores the need for a structured framework to facilitate harmonisation, presenting the first guideline framework and workflow. Additionally, it aims to assist researchers new to harmonisation by offering a guide matrix featuring examples from published literature, aiding in selecting appropriate integration models.

## Data Availability

All data produced in the present work are contained in the manuscript.

## List of abbreviations

AE: Autoencoders
AI: Artificial Intelligence
CFG: Context Free Grammar
CNN: Convolutional Neural Network
CT: Computed Tomography
DB: Dense Block
DNA: Deoxyribonucleic Acid
DNN: Deep Neural Networks
ECG: Electrocardiogram
EEG: Electroencephalogram
EMR: Electronic Medical Records
EN: Elasticnet
FCNN: Fuly Connected Neural Networks
GAT: Graph Attention Networks
GCN: Graph Convolutional Network
GLM: Generalized Linear Models
GWAS: Genome-wide Association Studies
HMLN: Heterogeneous Multi-layer Network
kNN: K-nearest Neighbours
LN: Lymph Node
LSTM: Long Short-term Memory
MA: Manifold Alignment
MKL: Multiple Kernel Learning
ML: Machine Learning
MRI: Magnetic Resonance Imaging
NLP: Natural Language Processing
NMF: Non-negative Matrix Factorisation
NMR: Nuclear Magnetic Resonance
PCA: Principal Component Analysis
PE: Pulmonary Embolism
PRISMA: Preferred Reporting Items for Systematic Reviews And Meta-analyses
RNA: Ribonucleic Acid
RWR: Random Walk with Restart
SNF: Similarity Network Fusion
SNP: Single Nucleotide Polymorphism
SSAE: Stacked Sparse Autoencoder
SVM: Support Vector Machines
VAE: Variational Autoencoders

## Ethical Approval (optional)

Not applicable.

## Consent for publication

Not applicable.

## Author’s Contributions

**Murali Aadhitya M S:** Data curation, Visualisation, Writing - original draft.

**Mithun Mitra:** Supervision, Validation, Writing - review & editing.

**Sonika Tyagi:** Conceptualisation, Data curation, Supervision, Writing - original draft, Writing - review & editing

## Acknowledgements

We thank Tyrone Chen for his suggestions regarding concepts of the work done. We thank Sheba Sanjay for her writing and editing suggestions.

